# Effect of Compulsory Funding on Post-hospital Neurorehabilitation Outcomes and Length of Stay following Stroke

**DOI:** 10.1101/2024.10.14.24315499

**Authors:** Jason T. Lew, Frank D. Lewis, Mel B. Glenn, Gordon J. Horn

## Abstract

**Background and Purpose:** The American Stroke Association recommends post-stroke rehabilitation for those who qualify and have access to this service. Continued services at the post-hospital residential and outpatient rehabilitation levels of care result in further functional gains after acquired brain injury, including stroke, even when provided beyond 6-12 months post onset. Access to care remains a barrier to rehabilitation including third-party limits with funding and coverage restrictions. This study measured outcomes comparing those with state legislated Compulsory Funding (CF) vs. Restricted Funding (RF) for post-hospital neurorehabilitation.

**Methods:** There were a total of 402 participants across 14 states included in the study. The CF group had 201 and the RF group had 201 participants. The Mayo-Portland Adaptability Inventory (MPAI-4) was selected as the primary dependent measure.

**Results:** Post-hospital rehabilitation length of stay (LOS) was significantly shorter in the CF group, t(400) = 1.72, p < 0.05 (Cohen’s d = 0.17). A Mixed 2×2 RM-MANCOVA revealed a significant main effect of time of testing, Pillai’s Trace f(1,398) = 402.6, p < 0.001; power to detect = 0.99, partial eta^2^ = 0.50. The analysis also revealed significant between groups main effect f(1,398) = 12.8, p< 0.001, power to detect = 0.94, partial eta^2^ = 0.031. The results of Bonferroni post-hoc pairwise comparisons are presented.

**Conclusions:** This study examined the differences in rehabilitation outcomes and LOS for persons receiving CF post-acute neurorehabilitation with individuals receiving services in other states with RF. While both groups improved, the state legislated access to care CF group started post-hospital neurorehabilitation without delay, discharged more quickly thereby reducing cost, with a lower level of disability (MPAI-4 Indices) at discharge compared to the RF group.

## 1. Introduction

Stroke is a leading cause of severe, long-term disability in the United States [1]. Every year, approximately 15 million people worldwide are affected by stroke with up to 50% of those surviving with permanent and chronic disability [2]. Following a stroke, The American Stroke Association recommends acute inpatient rehabilitation for those who medically qualify and have access to this service financially. The average acute inpatient rehabilitation lengths of stay for mild, moderate, and severe levels of impairment after stroke were 8.9, 13.9, and 22.2 days, respectively [3]. However, it is estimated that only about 70% of patients are functioning well enough to return home after completion of acute inpatient rehabilitation [4]. Implementation of early intensive rehabilitation can lead to improved, sustained functional gains [5, 6]. It has also been demonstrated that continued services at the post-acute residential and outpatient rehabilitation levels of care results in further functional gains after acquired brain injury, including stroke, even when provided beyond 6-12 months post onset [7, 8].

Frequently, access to care is a barrier to continued rehabilitation and recovery. This includes lack of third-party funding or limited coverage restricting rehabilitation therapies and/or availability of care near a person’s home when discharging from a designated stroke center. Prior research has suggested that disparities in post-stroke functional status are associated with insurance status [8]. However the cost savings from functional improvements and increased independence gained from improved access to early rehabilitation (acute and post-acute) more than offset the cost of services. This rehabilitation cost-offset has been demonstrated within average time frames ranging from 14 months to 5 years in those with acquired brain injury. The estimated overall average in lifetime cost savings is $1.67M [9].

A post-hospital outpatient program in California consisting of 6 hours of therapy services for 5 days a week was estimated to provide a lifetime savings of 1.2M for post-stroke patients with moderate to severe disability [10]. Similar cost benefits have been noted in the United Kingdom, with an average of $1.27M of lifetime savings among those with acquired brain injury who were enrolled in an interdisciplinary post-hospital program [11]. The savings were particularly pronounced in patients who received rehabilitation within one year of their injury. In addition, persons enrolled after one year of injury showed an economically significant difference compared to those who did not participate in a rehabilitation program. Cost savings have been found to be significant in those with chronic acquired brain injury up to 6 years post-injury with the implementation of post-hospital residential community integration programs, supporting the use of rehabilitation programs with chronic injuries [12]. Access to stroke neurorehabilitation can be varied demonstrated by prior research. Outcomes differ by access as well. For example, stroke survivors starting rehabilitation earlier with lower levels of independence, offset this cost in a shorter time period than those who start with higher levels of independence but have a delay with treatment [9].

While most states do not regulate the necessity of post-acute rehabilitation services, the state of Texas passed legislation in 1995 for compulsory funding focused on acute and post-acute acquired brain injury rehabilitation. However, this does not apply to all insurance policies within that state. The government sponsored Medicare program, and some larger third-party policies remain exempt from this legislation [13]. Currently, the covered programs include acute inpatient rehabilitation facilities as well as post-acute rehabilitation programs with neurorehabilitation and neurocognitive therapies. The post-acute rehabilitation programs include outpatient day treatment services, post-acute residential transition services, and community reintegration services. These post-acute rehabilitation programs may be implemented after discharge from acute inpatient rehabilitation, skilled nursing facilities, assisted living programs, or from home.

The aim of the current study was to evaluate post-acute rehabilitation efficacy and efficiency with reducing disability from admission to discharge among patients with stroke receiving services under the Texas Compulsory Funding (CF) legislation compared to patients who had Restricted Funding (RF) access based on third-party limitations. More specifically, the current research hypothesizes that stroke patients enrolled in a post-hospital residential rehabilitation program funded under the CF within the state of Texas will demonstrate greater improvement in physical and cognitive abilities than those enrolled in similar residential programs in states with RF (primary hypothesis) in the context of earlier admission and shorter length of stay (LOS). In addition, greater improvement of neurobehavioral adjustment and societal participation through applied skills use will also be demonstrated by those with CF as compared to RF (secondary hypotheses).

## 2. Methods

### 2.1 Subjects

Study participants were selected from a total sample of 753 consecutive admissions to post-hospital residential rehabilitation programs located across 14 states in the United States from 2012 through 2024. Of the 753 subjects with a stroke diagnosis, 590 persons met the criteria to participate. Within the possible 590 persons throughout the United States, 201 individuals were treated in states with RF services, while 394 individuals were treated within the CF group. In order to reduce random variability inherent in unequal sample sizes, 201 persons were randomly selected with SPSS from the 394 in the CF group. SPSS was used for data analysis including group comparisons as well.

The inclusion criteria for the study participants included a diagnosis of stroke, 18 years or older at the time of the neurological event, time from stroke onset to post-hospital admission of 28 days (i.e. chronicity) or longer, and participation in this level of neurorehabilitation at least 10 days or greater. The exclusion criteria included dual neurological diagnoses (e.g., other acquired brain injury) and persons participating in residential supported living programs without active treatment or in day treatment programs since the day environment and program vary considerably among subjects. Specific to this study, the only state with CF for post-hospital care was Texas. Therefore, the study participants in this group were from Texas only.

### 2.2 Measurement

The Mayo-Portland Adaptability Inventory (MPAI-4) is a measure that has been used extensively in post-hospital rehabilitation programs. It consists of 30 items with scores ranging from 0 (no disability) to 4 (severe impairment). The MPAI-4 contains three subscales: Ability Index, Adjustment Index, and Participation Index. The Mayo Portland was selected as the dependent measure due to its psychometric properties. Rasch analysis has been used extensively with the MPAI-4 to demonstrate item reliability, along with concurrent and construct validity [14, 15]. For detailed description of the MPAI-4 with psychometric properties, including Pearson interrater reliability coefficients, see the instrument manual [16]. Ratings on MPAI-4 Abilities, Adjustment and Participation Indices were converted to T-scores and comprised the primary dependent measures in the study. These are interval level measures of disability that are appropriate for use with parametric statistics.

### 2.3 Procedures

Each facility was part of a nationwide health network that is distributed in different regions of the country with the exception of the northwest. This distribution of facilities provides a reasonable representation of the United States demographically for post-stroke care. In addition, each facility specializes in post-hospital neurorehabilitation residential services for individuals with various neurological disease and conditions, including stroke. The neurorehabilitation program for all subjects provided physical (PT), occupational (OT), and speech therapy (SP) services 5 days per week, with 24/7 nursing support, weekly physician support, weekly case management, psychological counseling, and supervision. At this level, management of physical, cognitive, and communication deficits are part of the rehabilitation programming with the goal of returning home within the community.

Participant characteristics, including indicators of severity at the time of admission, are presented in Table 1.

**Table 1.** Participant Characteristics.

| Person Variables | CF Group | RF Group |
| --- | --- | --- |
| Age | 55.7 (9.8) | 51.9 (11.4) |
| Gender | 119 male, 82 female | 131 male, 70 female |
| Length of stay | 93.7 (72) | 121.8 (220) * |
| Chronicity | 116.6 (286) | 294.6 (808) ** |
| Severity of Injury |  |  |
| **Indices** | **Admission T-scores** | **Admission T-scores** |
| MPAI-4 Abilities | 57.6 (7.9) | 59.8 (10.9) |
| MPAI-4 Adjustment | 51.8 (7.1) | 57.9 (10.2) * |
| MPAI-4 Participation | 58.1 (9.5) | 59.5 (9.2) |

Note: Standard deviation indicated in parentheses. The Mayo Portland Adaptability Inventory – 4 (MPAI-4) T-scores (lower score = reduced disability). CF = Compulsory Funding. RF = Restricted Funding.
\* Significant at $p < 0.05$
\*\* Significant at $p < 0.01$

Subjects in the CF group were modestly older (55.7 vs 51.9 years), more female participants (41% vs 35%), less chronic (defined as the length of time from injury to post-hospital admission), with a shorter LOS (93.7 vs. 121.8 treatment days) than the RF group. Severity of injury at admission was measured by MPAI-4 T-scores (Abilities, Adjustment and Participation). The level of disability for both groups at the time of admission placed participants within the moderate to severe range indicating the need for post-hospital residential neurorehabilitation care with close supervision.

### 2.4 Statistical Analysis

The study hypothesis was tested using a Mixed 2×2 (group x time of testing) Repeated Measures Multivariate Analysis of Covariance (RM-MANCOVA) design. Significant RM-MANCOVA findings were followed with Bonferroni post-hoc tests for group comparisons. Post-hoc paired sample t-tests were used to evaluate differences from admission to discharge. Admission group differences for length of stay and chronicity were analyzed with independent sample t-tests. All analyses were performed using SPSS version 29.

## 3. Results

Length of stay in program and chronicity (length of time from injury onset to admission) can have a significant effect on outcome. Therefore, independent sample t-tests were performed to determine group equivalence on those variables prior to hypothesis testing. The differences were significant for LOS, t(400) = 1.72, p < 0.05 (Cohen’s d = 0.17), and chronicity, t(400) = 2.94, p < 0.01 (Cohen’s d = 0.29). Given the significant differences, both variables were entered as covariates in the analysis comparing outcomes for the two groups. The Mixed 2×2 RM-MANCOVA revealed a significant main effect of time of testing, Pillai’s Trace f(1,398) = 402.6, p < 0.001; power to detect = 0.99, partial eta^2^ = 0.50. Table 2 shows the results of post-hoc paired sample T-test.

**Table 2.** Results of Post-hoc paired sample t-test.

| **CF Group** | **MPAI-4 Indices** | **Admission** | **Discharge** | **Difference** | **t-value** | **Cohen's d Effect Size** |
| --- | --- | --- | --- | --- | --- | --- |
|  | Abilities | 57.56 | 50.14 | **7.42\*** | t(200) = 18.01 | 1.27** |
|  | Adjustment | 51.78 | 46.13 | **5.65\*** | t(200) = 11.12 | 0.78** |
|  | Participation | 58.11 | 50.20 | **7.91\*** | t(200) = 14.02 | 0.99** |
| **RF Group** | **MPAI-4 Indices** | **Admission** | **Discharge** | **Difference** | **t-value** | **Cohen's d Effect Size** |
|  | Abilities | 59.79 | 51.61 | **8.18\*** | t(200) = 15.09 | 1.06** |
|  | Adjustment | 57.87 | 50.42 | **7.45\*** | t(200) = 13.57 | 0.96** |
|  | Participation | 59.47 | 51.76 | **7.71\*** | t(200) = 14.30 | 1.00** |

Note: CF = Compulsory Funding, RF = Restricted Funding
\*Mean difference is significant at the $p < 0.001$ level.
\*\* Effect size = large effect.

Both groups showed significant improvement from admission to discharge on each of the three MPAI-4 indices. Large effect sizes were observed for each comparison.

The analysis also revealed significant between groups main effect f(1,398) = 12.8, p< 0.001, power to detect = 0.94, partial eta^2^ = 0.031. The results of Bonferroni post-hoc pairwise comparisons are presented in table 3.

**Table 3.**
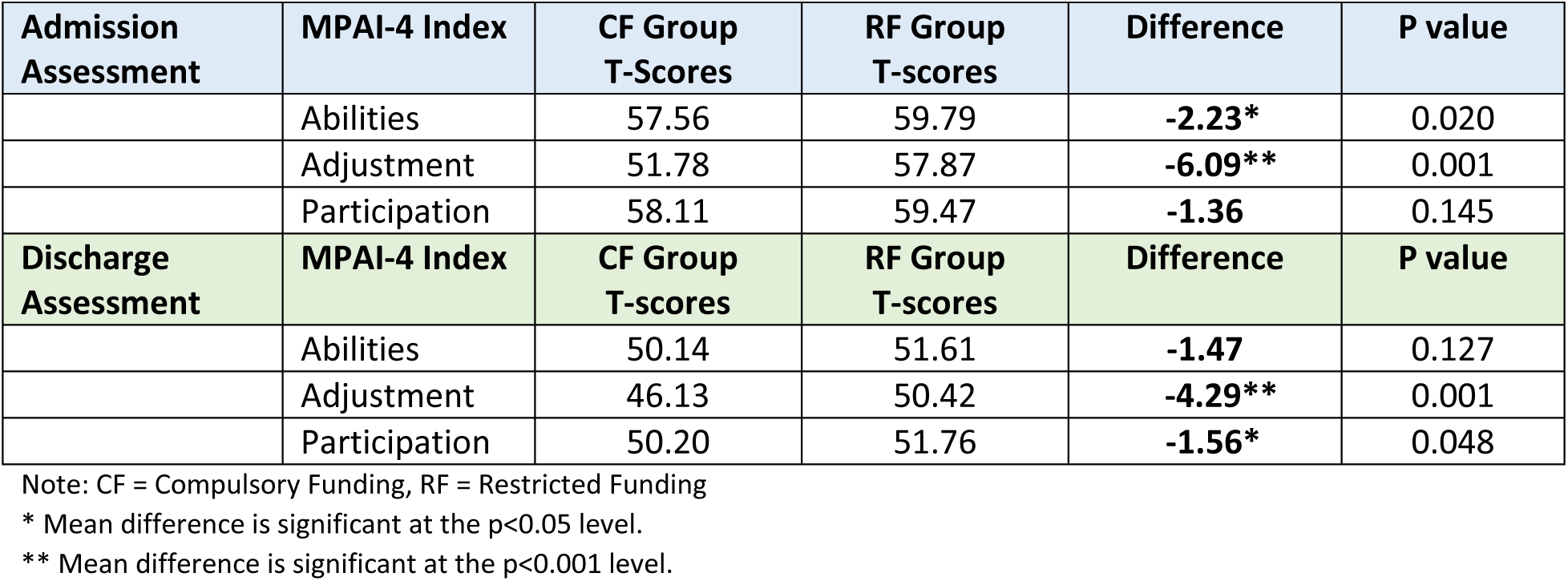
Bonferroni Post-hoc between group comparisons at Admission and Discharge for MPAI-4 T-scores.

At admission the CF group had significantly lower scores (i.e. less disability) on the Abilities Index (p <.05.). At discharge the CF group’s Ability scores were lower, but differences between the groups were not significant. Differences on the Adjustment Index were significant at both admission and discharge (p < 0.001), indicating less disability for CF participants on measures of neurobehavioral adjustment. The groups did not differ at admission on the Participation Index. However, differences were significant at discharge (p <.05). Individuals in the CF group experienced less disability with self-care and community participation skills following discharge.

## 4. Discussion

Neurological rehabilitation is complex from a financial and service delivery perspective. From the service delivery, there are many options ranging from inpatient rehabilitation facilities (e.g., acute rehabilitation care) to post-hospital residential programming focused on improving a person’s skills and abilities in a real-world setting prior to returning home for sustained and successful outcome.

Financially, limited legislation has been passed in Texas indicating “rehabilitation care cannot be denied if such services are medically necessary as a result of and related to an acquired brain injury including stroke” [17]. In addition, services do not have to be restricted to the hospital level of care, but can be provided in a community setting that is approved for rehabilitation.

This study examined the differences in rehabilitation outcomes and LOS for persons receiving CF post-acute neurorehabilitation in Texas with individuals receiving services in other states with funding limitations. The results indicate that while both groups improved, the State legislated access to care CF group started with less disability since access to care was without delay. This group also discharged more quickly with a lower level of disability (MPAI-4 Indices) compared to the RF group. The RF group demonstrated higher disability at the time of admission, and required a longer LOS before an acceptable discharge could be achieved. It is also notable that the RF group did not end their rehabilitation at the same level of reduced disability as the CF group. The statistically significant improvement in the RF group, despite the chronicity, supports prior research that there can be continued improvement in post-acute rehabilitation programs even with an extended length of time since injury [18].

Injury chronicity (time of stroke onset to post-acute neurorehabilitation admission) was different among the two groups as well, with the CF group having less time from injury onset to admission than the RF group. The CF group averaged 116.6 days post-stroke at the time of enrollment as compared to the RF group who averaged 294.6 days. Given the importance of early rehabilitation to maximize recovery, this significant difference in access to care may be reflected in the improved MPAI-4 scores at discharge in those patients who admitted to the rehabilitation program sooner [19]. However the interpretation of these results must be tempered since there was no information available regarding any possible interventions received between acute care and post-hospital rehabilitation admission.

Regarding length of time in post-hospital treatment, the CF group had a shorter LOS in the neurorehabilitation programs. On average, the length of stay was significantly different (p<0.05) for the CF group (93.7 days) as compared to the RF (121.8 days). The difference of 28 days in rehabilitation provides a substantial cost savings when entering post-acute rehabilitation as early as possible following acute care. The cost impact can be calculated by totaling the number of days in program then multiplying the daily cost of all services for a total cost savings. In addition to the shorter LOS, the CF group discharged at a lower level of disability.

Regarding adjustment or neurobehavioral effects of injury toward outcomes, the CF group was less behaviorally impaired than the RF group as evidenced by the MPAI-4 Adjustment Index for both admission and discharge. The Adjustment Index is used to evaluate mood and interpersonal interactions. This index includes ratings for anxiety, depression, irritability and/or aggression, symptom sensitivity, and social-behavioral awareness. The delay in starting post-acute rehabilitation may have led to maladaptive behavioral changes not identified in the CF group. The overall change from admission to discharge was larger for the RF group as well. This finding is consistent with improvement in emotional adjustment secondary to the structure and support of the neurorehabilitation milieu. Depression and anxiety after stroke have been associated with worse outcomes and lower quality of life. These programs also treat concomitant anxiety and/or depression thereby improving the MPAI-4 Adjustment Index important for post-stroke recovery [18, 20, and 21].

Societal participation evaluates applied skills that must be generalized to the community for successful outcome as measured by the Participation Index [18, 22]. At the time of admission, the two groups were not significantly different (p = 0.145, n.s.). There was a significant difference (p<0.05) at discharge with the CF group experiencing modestly less difficulty with applied skills such as cognitive and behavioral initiation, self-care, and home skills management. These are necessary components in measuring the level of community reintegration which is a frequent concern for those affected by a stroke. Improvement with these skills also reduces the burden of care by friends and families when discharged to home. Approximately 39-65% of people with stroke living in the community report difficulties reintegrating into community activities [23]. Prior research has also reported that the Participation Index is the least likely of the three Indices to return to non-impaired levels. Therefore, gains made within this Index produces a meaningful difference in disability reduction for persons after stroke [15, 24]. From the current study results, entering a post-acute neurorehabilitation program more quickly after a stroke may allow for increased gains in social participation upon discharge to home.

One unintended finding was the total number of persons available for the study. Twice as many study participants were available from Texas than the other 13 states in the study combined. At the time of this study, Texas was the only state with CF suggesting that post-stroke rehabilitation benefits from this legislation to increase access to specialized care following stroke. The end result is quicker admission for services with reduced length of stay and better outcomes than those with RF negatively impacting outcome. CF vs. RF also raised the question of level of impairment to qualify for care at the post-hospital level. The CF group, by definition of the legislation, did not require a specified level of impairment to justify the post-hospital residential neurorehabilitation as long as services were medically necessary. However, RF subjects may have required a higher level of disability based on external criteria, with emphasis on self-care impairment, to qualify for this level of service. This is often the case with third-party commercial policies.

There were limitations to the current study as well. First, post-discharge follow-up data was unavailable for this study to determine stability of gains. Post-discharge gains have previously been shown to be maintained at one-year follow up providing a measure of positive return on investment from neurorehabilitation care [25]. Second, there was no data to include where persons were coming from when obtaining post-hospital rehabilitation. The persons in the study mostly likely have been referred for services from acute care, but may also admit from home, skilled nursing or long-term care. Third, stroke laterality was not obtained. Those with aphasia may have a greater level of impairment due to the language disturbance. A follow up study may replicate the methods and results with a traumatic brain injury sample.

## Data Availability

Due to the nature of the research (proprietary data), the data in the manuscript is not available.

## Acknowledgements

None

---

**Figure 1.**
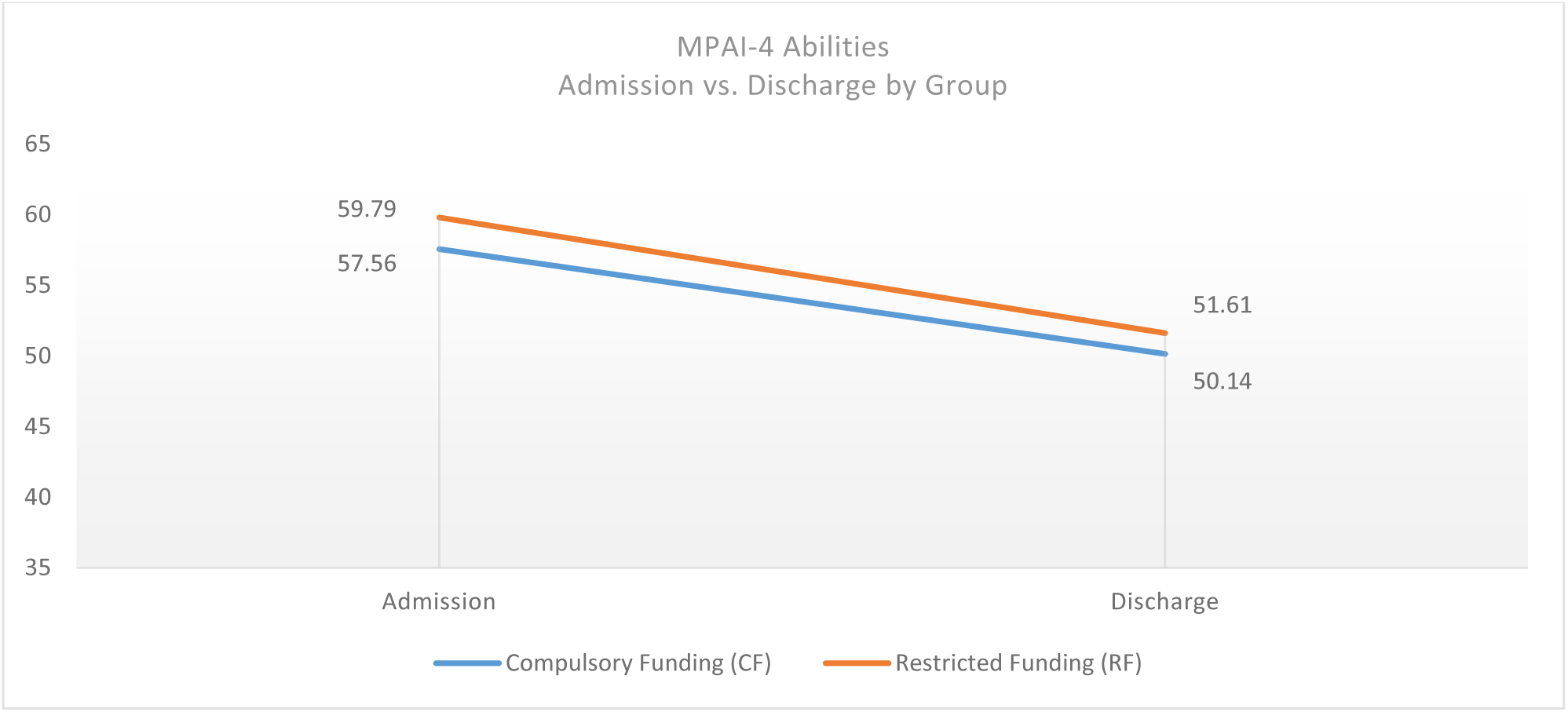
MPAI-4 Abilities Index – Admission to Discharge disability reduction.

**Figure 2:**
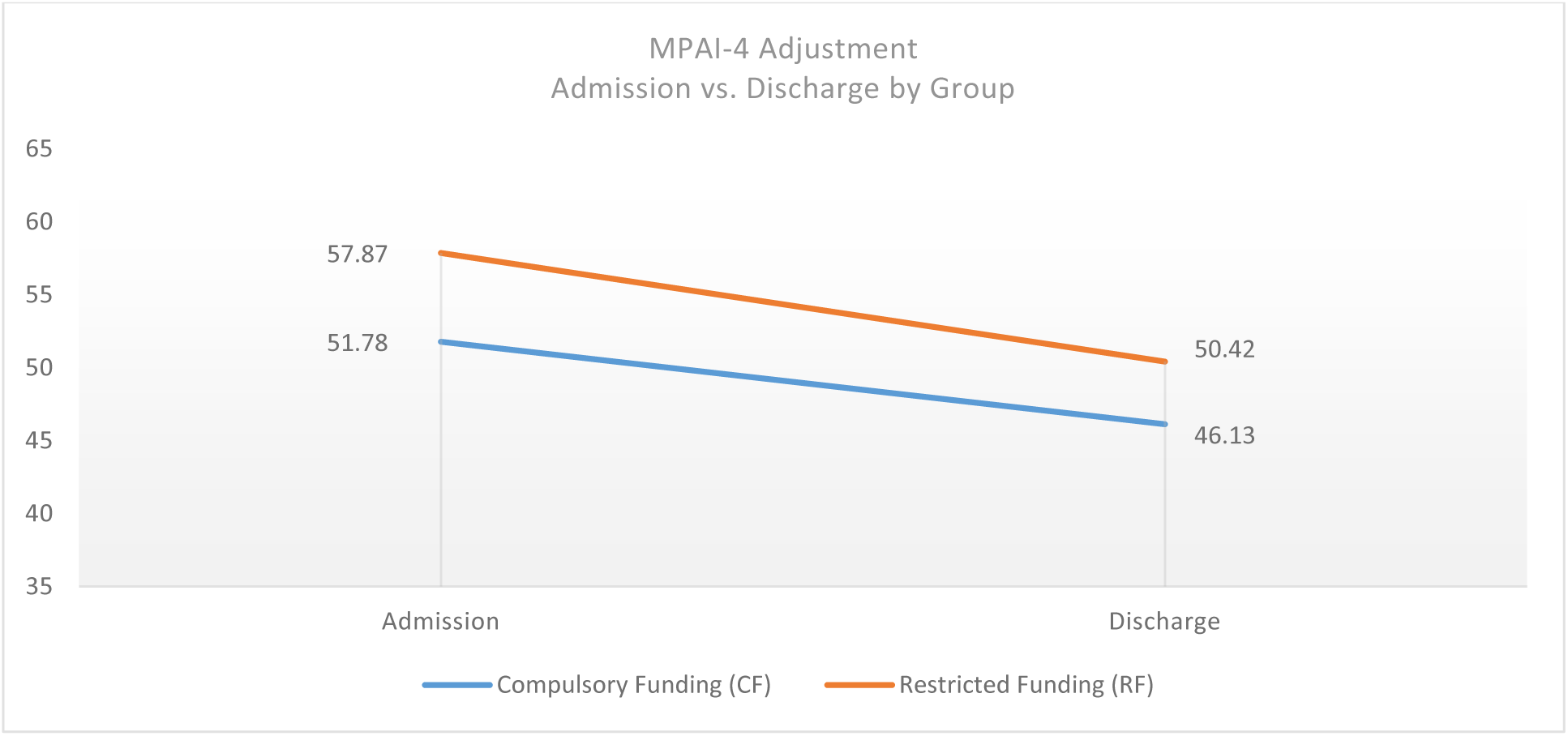
MPAI-4 Adjustment Index – Admission to Discharge disability reduction.

**Figure 3.**
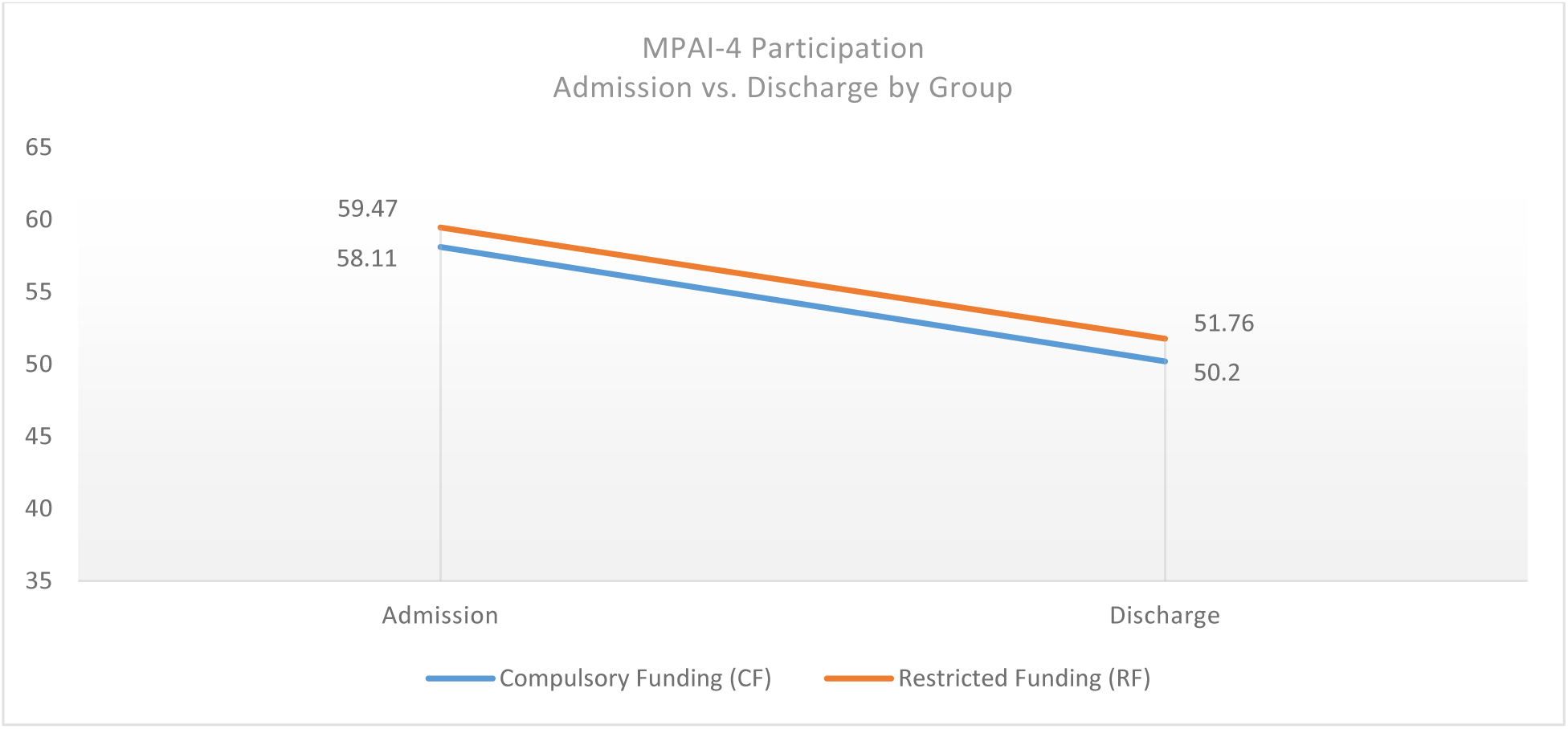
MPAI-4 Participation Index – Admission to Discharge disability reduction.

